# “We saw them as stories”- Understanding how multidisciplinary case review contributes to quality improvement in trauma care

**DOI:** 10.1101/2025.08.08.25333298

**Authors:** Johanna Berg, Helle Mölsted Alvesson, Martin Gerdin Wärnberg, Nobhojit Roy, Ulf Ekelund, Siddarth David

## Abstract

The effects of quality improvement interventions in healthcare are mixed, and the mechanisms through which they mediate their effect remain poorly understood. Quality improvement methods often rest on implicit assumptions of predictability, linear causality, and standardisation. Health systems are increasingly recognised as complex adaptive systems, where outcomes emerge through adaptation, self-organisation and non-linear interactions. Trauma, defined as injury and the body’s response, is the leading global contributor to quality-related mortality. As trauma care involves a wide range of injuries, fragmentation across departments and time, and the need to coordinate care without complete information, it is well suited for examining how improvement unfolds in a complex adaptive system. Although trauma QI programs are widely used, particularly in high-income countries, how they contribute to improved care remains unclear.

We examined how multidisciplinary case review supported improvement in two tertiary hospitals in urban India, as part of a trauma QI program. Using reflexive thematic analysis informed by complexity theory, we identified four mechanisms: (1) system-level situational awareness, which helped providers see the system as a whole and understand how their actions affected care over time; (2) shared understanding, which created a common narrative that reduced blame and enabled collaborative problem-solving; (3) navigating systems of power, which involved working within constraints, using informal influence, and advocating for change where needed; and (4) ethical sensitisation, which emerged as providers saw the patient’s full story and experienced a personal, emotional recognition that change was necessary to avoid harm. We suggest that improvement may be understood as an emergent property in a complex adaptive system, and conditions that support the emergence of the mechanisms we identified may in turn lead to improvement in outcomes.

## Background

Quality improvement can be described as a systematic, continuous approach to improving healthcare processes and outcomes through data-driven change and collaborative problem-solving^1,2^. The concept of quality improvement has its roots in industrial manufacturing, where methods were developed to improve efficiency, reduce errors, and ensure consistency in large-scale production. The methods used were based on a reductionist view of systems as collections optimisable parts where improving each component would generate better overall performance, as such, assuming a predictable, linear relationship between cause and effect. This perspective also gave rise to a top-down managerial style, where control and decision-making were centralised. Over time, techniques evolved to manage increasingly complicated processes, especially in high-reliability areas of manufacturing. Despite differences in application when used in other areas, such as health-care, many quality improvement approaches continue to rest on implicit assumptions of predictability, standardisation, and controllability^2^.

Health care, and health systems, are increasingly being recognised as a complex adaptive system. A complex adaptive system can be defined as a collection of individual agents that have the freedom to act in ways that are not always predictable, and whose actions are interconnected such that one agent’s actions change the context for other agents^3^. Hence, it consists of multiple interacting agents whose behaviours are shaped by feedback, local context, and adaptation over time. Because interactions are dynamic and interdependent, complex systems are inherently non-linear and the relationship between cause and effect is not proportional. As actors in the system continuously respond to another, patterns and outcomes emerge in the system. These features make uncertainty a defining property of complex adaptive systems where the future state of the system cannot be fully predicted in advance, even when all components are known^4–6^. In essence, health care is not a rigid, predictable system; it is a complex adaptive system requiring constant clinical reasoning, adaptability, and patient-centred decision-making. The ability to provide individualised, high-quality care is often dependent on contextual adjustments that fall outside the boundaries of predefined protocols^4,7,8^.

Despite the widespread implementation of quality improvement initiatives over the past decades, evidence of their effectiveness remains mixed. Systematic reviews have highlighted both the heterogeneity of interventions and variability in outcome selection as barriers to meaningful synthesis and comparison across studies^9^. Reported effects vary considerably, with some reviews suggesting that between 50% and 70% of high-quality studies demonstrate improvements in outcomes^10^. One explanation offered is poor adherence to defined quality improvement protocols or insufficient implementation support^10^. However, the field has also been criticised for being under-theorised, with limited understanding of how interventions mediate their effects, making it difficult to determine why some programs succeed while others do not^11^. In response, there have been increasing calls for mixed-methods research and theory-informed evaluation approaches to better explain *how* quality improvement contributes to improved care^1,12^.

Trauma is defined as physical injury caused by external forces, along with the body’s associated physiological response. It accounts for more than four million deaths annually and is the leading cause of death among people aged 10–29 years. Most trauma deaths occur in low- and middle-income countries, and it is estimated that over two million lives could be saved each year if outcomes in these settings matched those in high-income countries^13^. Trauma is the leading contributor to quality-related mortality globally, making it a critical target for improvement^14^. Delivering trauma care spans prevention, prehospital care, emergency and hospital treatment, and rehabilitation. The medical management is characterised by high clinical variability, and presentations range from isolated fractures to complicated multisystem injuries and physiological complications. Care is often time critical, and decisions must be made under high uncertainty and with incomplete information. Because no protocol can anticipate every clinical scenario, teams must continuously reassess physiology, interpret findings, and adapt interventions in real time. These features, the dynamic interaction between actors, adaptation in handling uncertainty, and emergent behaviours, make trauma care an example of a complex adaptive system^5,8^. Early on, trauma practitioners recognised the need for systems approaches to improve patient outcomes, and the field of trauma care was early to adopt structured system approaches, including regional trauma systems and quality improvement programmes based on data collection, audit filters, and multidisciplinary case review^15,16^. Still, the effectiveness of these programmes, and the mechanisms through which they may contribute to improved outcomes, remain poorly understood. To overcome the global quality gap for trauma care, understanding how these programs lead to improved care and patient outcomes is essential.

This study aimed to examine how patterns emerging during multidisciplinary case review may act as mechanisms that contribute to improvement in a complex health system.

## Methods

### Study design

We conducted an interview study during the final phase of a five year controlled interrupted time series trial evaluating a trauma quality improvement program. The trial, the Trauma Audit Filter Trial (TAFT, ClinicalTrials.gov: NCT03235388), was conducted at four tertiary care hospitals in urban India. After a one year observation period two hospitals implemented a trauma quality improvement program, while the other two continued standard care. The program consisted of three main components: prospective data collection, identification of deviations from clinical standards using audit filters, and regular multidisciplinary review meetings. Audit filters are predefined statements describing expected care processes in clinical situations, such as securing an airway in patients who are unconscious. A multidisciplinary review board was established at each intervention hospital, consisting of senior healthcare staff from departments including emergency, general surgery, anaesthesia, orthopaedics, neurosurgery, radiology, and plastic surgery. The local principal investigator at each site coordinated the recruitment of board members. Each hospital selected a set of locally adapted audit filters through an anonymous online Delphi process^17^. Trained project officers prospectively identified trauma patients and recorded whether any audit filters had been flagged and collected outcomes and interventions during the hospital stay. These flagged cases were compiled into structured reports where the trajectory of care during the hospital stay could be followed for each patient. Both summaries of flagged cases, and individual cases of interest were presented at regular multidisciplinary meetings. During these meetings, the board identified areas for improvement and proposed actions to address them. After the initial implementation phase of six months, the meetings were conducted independently by the hospital teams without involvement from the research group^17^. We used reflexive thematic analysis^18^, with a theory informed interpretation guided by complexity theory. We explored how patterns of interaction and adaptation emerged through multidisciplinary case review, and how these may act as mechanisms that support improved care in a complex adaptive system.

### Participants

All personnel who participated in the interdisciplinary case review meetings during the trial were invited to take part. This included senior physicians, surgeons, nurses, and project officers. The project officers, who had no clinical role, were responsible for prospective data collection. All participants were aware of the goals and rationale of the TAFT trial but were not informed of the quantitative results at the time of the interviews. In total, 23 individuals participated in the interdisciplinary case reviews at both hospitals. One unfortunately passed away during the trial, so 22 were approached and 20, 15 male and 5 female, agreed to be interviewed (Table 1). Recruitment was conducted via email invitations, and all participants provided written informed consent before their interviews. Interviews were conducted in person at the hospitals where participants were employed, except for the project officers, who were interviewed in an external setting. One interview was conducted via Zoom at the participant’s request. While most interviews were conducted privately, occasional workplace interruptions occurred.

**Table 1:** Characteristics of participants by department.

| Specialty / Department | Number of participants | Roles included | Experience |
| --- | --- | --- | --- |
| General surgery | 6 | Professors, Associate Professors | 10–20 years |
| Orthopedic and neurosurgery | 4 | Professors | >10 years |
| Emergency and anaesthesia | 4 | Senior Nurse, Senior Physician, Associate Professor | 5–10 years |
| Other specialties (Radiology, ENT, Plastic) | 4 | Professors, Assistant Professor | 5–20 years |
| Research team, data collection | 2 | Project Officer | 5–10 years |

### Data collection

A semi-structured interview guide was developed in collaboration with the TAFT research team. The guide focused on participants’ experiences with the audit filter process and interdisciplinary case reviews, and was informed by the clinical and contextual knowledge of the research team, as well as the aim to understand how the intervention worked. To support this, the guide was structured around three domains: the existing state of trauma care, the experience of implementing the intervention, and its perceived effects. It was also designed to capture participants’ interpretations of the process and to prompt reflection on how the intervention may have influenced care. The guide was not piloted before data collection but was refined iteratively during early interviews.

Interviews were audio-recorded, supplemented by field notes, and conducted over a two-week period. All interviews were conducted by both JB and SD, where we took turns and for each interview, one led the discussion while the other took notes and posed follow-up questions to explore key topics further. Interviews lasted 30 to 45 minutes. Transcription was performed by JB. Ten interviews were transcribed using NoScribe, an open-source, AI-based transcription tool that runs locally to preserve data privacy and confidentiality^19^. All automated transcripts were manually verified for accuracy.

### Data analysis

The analysis was inductive, based on open coding with no pre-specified framework. Once familiar with the transcripts, both researchers jointly coded two interviews before proceeding independently, with JB coding 9 and SD coding 8 of the remaining interviews. We did not stratify responses by professional role or hospital site, as our focus was on identifying emerging patterns that could be mechanisms for improvement. Codes were iteratively refined through regular discussion. Our approach prioritised interpretive depth over consensus, with codes and candidate themes developed and adjusted throughout the process. While the initial coding process was inductive, we used a complexity-informed lens to guide our interpretation. This allowed us to focus not on linear cause and effect, but to actively look for emergence and, in particular, patterns that could support improvement. Quotes were lightly edited for clarity and readability, with filler words and hesitations removed where they did not affect meaning. All edits preserved the tone and intent of the speaker.

The analysis was conducted by two researchers with complementary perspectives. JB, a Swedish emergency medicine physician and PhD candidate, contributed a clinical viewpoint but was an outsider to the study context. SD, a public health scientist from India with extensive experience in trauma research, provided contextual and systemic insight. Both researchers maintained reflexive notes to document assumptions and interpretive decisions. Their combined clinical and social science backgrounds and expertise shaped the perspective in the analysis and writing of this paper. While both JB and SD were core members of the TAFT trial team, not all coauthors were involved in the design or implementation of the intervention. Preliminary interpretations and findings were discussed among coauthors.

We used Taguette^20^ on a secure research server, accessible via VPN, for collaborative coding. De-identified codes and quotes were then exported to Google Sheets, which was used collaboratively to structure and organise themes. To protect participant anonymity, specialties are grouped in the participant table and not separated by hospital. For a small number of quotes that are more sensitive, no participant number is provided. Ethical approval for this study was obtained as an amendment to the existing ethical approvals for the TAFT project, granted by the local ethics committees at each participating hospital: MAMC (F.1/EC/MAMC/92/04/2022/No421, dated 07/11/2022) and SSKM (IPGME&R/IEC/2022/517, dated 14/11/2022).

## Results

We identified four emergent mechanisms through which multidisciplinary case review contributed to improvement: providers developed system-level situational awareness, an increased understanding of their role in the fragmented system they work in; they got a shared understanding across departmental boundaries that reduced blame and enabled collaboration; they learned to navigate hierarchical structures to initiate change and, lastly, they became sensitised to the consequences of their actions, creating a strong personal ethical motivation to improve.

### Theme 1 - System-level situational awareness: Making the fragmented system visible

Trauma care was described as fragmented, requiring coordination across departments and disciplines that often operated in silos. Providers typically focused on their own departmental priorities, which limited opportunities for collaboration or shared learning.

> *“That is the whole issue, because when there are different departments, everybody is looking at the priority of their department […] So everybody is focusing on their own work, thinking, ‘I’ve got so much work in my department, this is not a priority*.*’”* – Participant 6

Rotations of junior doctors and nurses further disrupted continuity and learning. Participants noted that training efforts were often short-lived, as staff turnover limited the retention of knowledge.

> *“We had put posters in the emergency so that the Glasgow Coma Scale (GCS) is easier to do. We had put up other posters, ABCDE, this is to be done. Initially, they made a difference […] and then the people who were trained in it left*.*”* – Participant 2

Specialisation deepened this fragmentation. Providers reported a reluctance to take responsibility for general trauma care, as their professional focus was increasingly narrowed to their own subfield.

> *“I find today it is the self-motivation, your commitment to society and to healthcare, that is missing. People are getting more into their own fields. One reason is that every field has become super specialized. You need to put so much more effort into your own area that this[trauma care], which is a general field, gets left behind. They do not think it is important. […] Even today, postgraduate students join orthopedics and say, ‘I want to be an arthroplasty surgeon, I want to be a spine surgeon*.*’ Basics, no one. That is why they are just focused on super specialization*.*”* – Participant 6

Being exposed to the patient care narrative, the story from admission to discharge, helped overcome the fragmentation of care. Participants could see how their own actions affected later parts of the patient’s care and helped them develop a system-level situational awareness: an understanding of how their actions, and those of others, interacted within the system.

> *“After the patient had been moved out from the ER, they gave us a picture of what happened to the patient. We got that feedback. We saw what was lacking on the ER side, what was lacking on the referral side, and in the last place the patient went. Getting that overall picture was very important for us. When we got that picture, we were able to analyze what lacuna we had, or what good work we were also doing, because minus and plus both are required. So we would go and encourage our residents: this is a good thing you have done, so you should do it more. And this is something that was lacking and should be incorporated*.*”* – Participant 3

The ability to overcome fragmentation, and to see the effects of the system in action, was made possible because of the ability to follow the patient’s journey throughout the hospital stay. And the emergence of system level awareness for participants became key to understand how and what to improve.

### Theme 2: Shared understanding: Reducing blame and creating a common cause

This system-level awareness extended beyond individuals. Seeing the same narrative of what happened to the patient appeared to strengthen a shared understanding of the problem, which in turn supported a sense of collective responsibility and helped reduce the tendency to blame others.

> *“The meetings provided a platform where people could sit and they could nod in agreement that, yes, we are together*.*”* –Participant 9

The shared situational awareness also created a foundation to collaborate and find interdepartmental solutions to identified problems. These solutions, which would be difficult to identify from outside this process, were derived from the knowledge, empirical experience, and local contextual understanding of the group.

> *“We learned a lot about how different departments work and how trauma care is a multi-specialty thing. That helped us pitch problems and solutions in a different way. For example, the emergency department did not realize the value of early E-FAST [Extended Focused Assessment with Sonography for Trauma] and ultrasound. After these meetings, they understood the problem — that GCS and respiratory rate were not being measured. They had only seen a snapshot at entry, not the outcome. We had seen the outcome, not the entry. Now everyone is sitting together and seeing the whole picture*.*”* – Participant 7

Participants also described how fear of judgement, both from the actors within the system, and from external actors, affected their capacity to work for improvement. Public and political scrutiny could create pressure to hide flaws, and within the system, blame created an atmosphere of fear. When providers felt unsafe, it became difficult to acknowledge gaps in their own practice.

> *“There is a fear in the government sector that if we pick out things, there will be punishment from the bureaucracy or from the political class. Once people are assured that this is not about punitive action but about improving the system, they will start getting involved. […] The resident or the medical officer on the floor also feels, how should I say, petrified that they will be judged, when judgment should not be happening in these situations. There are already a lot of people to judge us: the media, the public. If we judge ourselves, we cannot improve the situation. We have to be non-judgmental*.*”* – Participant 9

Review meetings and interdepartmental interactions could either reinforce fear or help counter it. The meetings appeared to support a culture of learning and mutual support rather than blame. Participants also emphasised the need for managerial involvement, mainly to ensure that leaders understood the same challenges and shared the same view of the system in order to support meaningful change.

> *“The positive thing about these meetings, because we do tend to have such meetings otherwise too, interdepartmental meetings. In those interdepartmental meetings, there is a lot of bickering and a lot of finger-pointing at others, like your department did this. I think the people in this trial were curated in a way that they had a positive mindset. They were encouraging and teaching the others: okay, you did this, but this can be done in that way. So the way a thing is conveyed, that was very important*.*”* – Participant 9

Having a shared view of what happened, how, and why created a sense of togetherness. Even when the discussions were about difficult events, focusing on what could be changed made the meetings feel constructive. Participants described this as something they looked forward to, an opportunity to reflect, connect with colleagues, and see that their efforts were leading to change.

> *“I personally used to look forward to them [the review meetings]. I enjoyed it. And I do not know if I should say that. But others also mentioned during the meetings that they looked forward to it every month. Yes, there were discussions about bad things, because mostly it was about why something went wrong. But still, having that cup of tea together and talking about what can be changed, everybody, and even I personally, felt very happy and positive about it. At least there was one hour where we would talk about solutions*.*”“* – Participant 7

Seeing the same story, narrative of the patients care in the hospital helped practitioners gain a common understanding and shared responsibility, which reduced blame and helped them identify collaborative solutions.

### Theme 3: Navigating systems of power: Acting within the constraints and possibilities revealed by system awareness

As system-level awareness emerged through multidisciplinary case review, participants not only recognised where care could be improved but also began to understand what was required to act on that knowledge. This included identifying what lay within their own control and what required advocacy in other parts of the hierarchical system they worked in.

A divide between generalists and specialists shaped how expertise was perceived, with specialization often seen as superior to broad clinical knowledge. This implicit hierarchy affected how responsibility was distributed. Some specialists positioned themselves as gatekeepers of knowledge and decision-making, while generalists or other providers were sometimes assumed to lack the necessary skills to manage certain conditions.

> *“ First point of contact must not be with the MBBS person. They make a mess of the head. Doesn’t understand the head injuries. Basic information is lacking in them*.*”* – Participant

> *“The sisters don’t play a major role in the management of the patient. They follow whatever we tell them*.*”* – Participant

These were not limitations of generalist providers, junior doctors, or nurses, but a result of how hierarchy structured professional interactions and learning. When providers saw the trajectory of patient care rather than just their own role, they recognised the need and importance of interdisciplinary collaboration. The solutions and training activities extended beyond doctors, identifying new ways for staff to contribute. When they did, it also became a way to empower staff, and they would volunteer to help when they identified a need and knew they had the skills to do so.

> *“We used to train our ward boys in CPR because we would fall short of manpower. At least chest compression—they should be very well versed with that. […] They would volunteer and say, ‘Let us do this and you take care of the complicated stuff*.*’ I think that made a huge impact*. – Participant 9

Constraints like understaffing and high patient volume led providers to adapt workflows and redistribute responsibilities. These changes were driven by senior staff using their local knowledge to find feasible solutions. This included sharing clinical expertise beyond speciality boundaries, allowing others to take on new tasks, and reorganising triage and supervision. These adaptations show how proximity to the work enabled adaptive responses.

> *“We are understaffed, we are two people who are running the emergency radiology services for the entire hospital and if they have to move around in the hospital or one has to be posted in the causality to be able to provide point of care ultrasound then it grossly reduces the manpower in the emergency services so*.. *Hence I also highlighted the importance of teaching the non-radiologists, the ability to provide point of care ultrasound*.*”* – Participant 3

> *“We have started categorizing the people needing imaging urgently. Considering the amount of patients, we always have a line of trolleys 24h outside of our CT… So we have moved the people who look sick or with a lower GCS ahead in the line so they can be imaged faster… We have the technician and the non-medical staff who are able to do that. We have one senior resident and two junior residents on duty, so one of the junior residents is supervising*.*”* – Participant 3

Formal mandates were not described as the main drivers of change. Instead, senior providers used their informal authority and professional standing to shape practice in their own departments. When senior staff led by example, junior providers were more likely to follow.

> *“Nobody gave us that okay from now on everything about trauma you people will decide. But from inside, if there were three people from surgery department, a few from emergency, then they went back to their departments and started making those changes in their work*.*”* – Participant 7

> *“If you see that their faculty is doing it regularly, I think that is the difference*.*”* – Participant 13

In contrast, there was also a sense of resignation to formal mandates, along with recognition that certain changes could not be achieved without administrative instruction. At the same time, the tension between mandating participation in the review board versus recruiting and empowering people was clearly highlighted. Mandates risked leading to disengagement and a loss of the crucial sense of personal and shared responsibility, ultimately rendering them ineffective as tools for improvement.

> *“Unless you mandate it, things never get implemented. So somewhere, you know, we have that colonial thought, you know, somebody is going to tell us how to do it*.*”* – Participant 13.

> *“You have to be careful when mandating something to senior faculty. If is presented as a discussion, let’s say an medical director said, ‘We need to do this, there are problems, trauma care isn’t good, and I think you’re the best people to lead this’, then it might still work. But if it’s just, ‘Give me the information and I’ll decide whether to enforce it*,*’ that doesn’t help. If you create an empowered committee, carefully select the members, and give them the authority to make changes, then yes, it can work. But if you just issue an order, without consultation, without discussion, and without giving them any power… and most committees are like that—nothing happens*.*”* – Participant 7

Lastly, hierarchy had a significant impact on how change could be achieved. The group initiating change needed to have enough mandate and seniority within the hierarchical system to be able to make it happen.

> *“We should target intermediate people, and that too through the chief. If you pick up a junior chief from the department, nobody listens to you”*. – Participant 19

The emergent mechanism seems to be the capacity to navigate the constraints of a fragmented, hierarchical system and find viable paths for action by recognising your own position and place of power. This included making changes within your reach, empowering colleagues around you, leading by example, and advocating upwards when necessary. The tension between mandate and self-driven engagement was central. While formal mandates could create structure, they also risked disengagement and, at worst, may serve as a way to deflect personal responsibility.

### Theme 4 - Ethical sensitization: An emergent response to seeing the consequences of care

When providers saw the full story of a patient’s care, and their place in the hierarchical system in which they work, it became clear how their own actions had shaped the outcome. This recognition felt personal and emotional. Participants described being “jolted” by the realisation that something they had done, or not done, had contributed to the outcomes and care of the patient. This process, described as sensitization, created a strong sense of personal responsibility.

> *“We, as clinicians, sometimes just go into auto mode without realizing how our actions impact the patient’s life. So sometimes we really need to be jolted, to realize, like, look, this is the change we need to make in the way we work. This is the data that has been generated, and if we do this, this is how things will change. So unless the person [referring to the provider] actually knows about it, there wouldn’t be a readiness to learn or to change their ways of work*.*”* – Participant 3

The experience of being “jolted” as a provider came from recognizing the tangible effects of one’s actions. There is a personal, ethical, and morally driven motivation that arises from this sensitization, the notion that the patient becomes more than just a body treated for a short time. This form of sensitization is not externally imposed, but internally felt, rooted in a personal ethical response to recognising the patient as a person, and oneself as part of their story. The capacity to act and change depending on these insights becomes personally imperative, and having the power to do act on these insights is key.

> *“The data tells a story, and the good part was that we were not bothered about statistics, like okay, you analyzed five patients and these, these, these were the difficulties. We saw them as stories also. So, when you see a person or you see a patient as a person, you tend to relate to that person, and that emotional bond sort of comes, and you tend to be more sensitive towards your actions or towards the missing of your actions, or both of them*.*”* – Participant 9

> *“I think when we made them [referring to residents] realize that if you did a particular thing, suppose if you recorded the GCS [Glasgow Coma Scale], it would have made the neurosurgeon’s job easy. Or if you had sent the blood group for cross-matching right away, then the patient could have, you know, could have got easy and early access to blood, which would have saved his life. So, the realization that these small things could have made changes to the patient’s outcome actually changed the resident’s behaviour. And that behaviour change, when it comes from the heart, I think it gets incorporated in the clinical practice*.*”* – Participant 9

This emerging pattern of sensitization, reinforced by system-level awareness, a sense of shared understanding, and recognition of one’s role and power within the system— represents a mechanism through which providers in a complex adaptive system are motivated to improve care. It shifts improvement from a task to a personal, relational responsibility grounded in meaning and ethics.

## Discussion

### Understanding the system through patient care narratives

Improving quality of care is about learning from empirical knowledge continuously. The fragmented nature of trauma care, both over time and across different departments and providers, makes it challenging for providers to understand the system they work in and learn from the care process. Exposing providers to the patient care narrative, the story of what happens from admission to hospital to the end of care, helps them gain a system level awareness of their own place and role within the system. When sharing the same narrative, they also create a common understanding which reduces blame. And as individuals, this allows them to identify changes within their power, or identify where to advocate for these changes in the hierarchical system they work in. It becomes ethically challenging not to act on things you identify within your power, as the alternative may cause harm. This process is sensitization, and it motivates providers to initiate change.

### Mechanisms that support improvement in complex systems

The identified mechanisms, system level situational awareness, shared understanding, navigating systems of power, and sensitisation, appear to emerge during well facilitated multidisciplinary case review. They align closely with theoretical concepts in complexity science, particularly how agents interact and generate sense making: the capacity to self organise and adapt their actions so that one agent will change their actions for others.^4^ To adapt effectively, providers need system knowledge, an understanding of what other actors or departments will do, what they are capable of, and how one’s own actions affect subsequent steps. This awareness is essential in a complex adaptive system, enabling providers to self-organise, adjust, and optimise decisions for themselves and their collaborators. System-level situational awareness and shared understanding are both forms of sense making: the process through which agents develop a shared interpretation of their experiences, a capacity linked to improved patient outcomes.^21^ Gaining system level awareness is the foundation that allows participants to identify areas of improvement and understand how that may be achieved.

The third mechanism, navigating systems of power, connects to two important aspects of complexity. The first concerns how to manage in a complex system, where leadership needs to be distributed to allow for self organisation more than top down approaches. This tension between formal mandates and self organisation was expressed by participants, who highlighted the duality of needing mandates for some things while at the same time acting without them for others. Navigating systems of power also relates to the dimension of human values in a complex adaptive system. Greenhalgh and colleagues describe how value perceptions about who holds expertise, who is allowed to decide, and what constitutes good care are implicitly part of all complex systems^22^. This was clearly reflected in our study, where participants actively used their gained system level awareness to navigate the hierarchical system around them, either to make changes or to advocate for change, depending on who held the power to do so.

The fourth mechanism, sensitisation, concerns a personal and ethical sense of awareness. It occurred when providers were emotionally jolted by recognising how their actions had directly shaped patient outcomes. These moments are described as emotionally powerful realisations where they described seeing the patient not just as a clinical case, but as a person, and recognising themselves as part of that person’s story. This recognition made it ethically difficult not to act, because the alternative might lead to harm. This reflects what Greenhalgh and colleagues argue: in complex systems, values are not abstract rules applied from outside, but are expressed and understood through narrative, emotion, and embodied experience^22^. Sensitisation captures this situated ethical awareness, where providers recognise how their actions relate to others and to the outcomes.

### Improvement in complex systems: Uncertainty, learning, and adaptation

In essence, medicine is a fundamentally uncertain applied science, and working within uncertainty is a reality of clinical work. This requires different approaches to clinical decision-making, drawing on formal evidence, physiological reasoning, empirical knowledge, and an understanding of who the patient is. In trauma care, this is particularly visible, as the care of a multiply injured patient spans several specialities and departments. The capacity to continuously learn from care narratives, and to develop empirically grounded knowledge across departments, may be key to improving care, particularly when care is highly unpredictable or uncertain. In such situations decisions must be made through experience and thoughtful clinical reasoning. As Sturmberg and Martin argue, clinical expertise emerges through repeated encounters with uncertainty, and improvement depends on recognising meaningful patterns in practice and building systems that support this kind of experiential learning^23^.

As complex adaptive systems constantly change and evolve, learning must be continuous. Improving care involves ongoing learning among providers, not only through new research, but through everyday, situated experiences of caring for patients. This kind of learning is empirical and collaborative, it involves understanding how care can be improved within the specific system in which providers work. Knowledge, in this view, is also generated inside the system, as providers collectively learn how to make good decisions in situations where formal evidence may be limited. Learning Health System frameworks relate to this, but are sometimes interpreted through a linear model in which knowledge is produced externally and then integrated into clinical practice. Recognising that important empirical and systems knowledge emerges from within the health system itself may help us better understand what kinds of structures and processes are needed to support continuous, practice-based learning^24^.

The tension between reductionist quality improvement methods and the realities of work in complex adaptive systems may create a misalignment between the methods used and the level of complexity where they are applied. Such approaches may be effective for more predictable problems or in settings with lower complexity, but are less suited to situations where complexity is high. This may be one reason why some quality improvement programmes succeed while others do not^25,26^. Overly technical interpretations of complexity can also risk overlooking the human dimensions of care, such as values, relationships, and ethical judgement, which influence how people act in real systems^27^. Recognising these tensions may help shift the design and support of quality improvement toward approaches that foster adaptive learning, relational reflection, and responsiveness to the dynamics of complex systems.

In this perspective, improvement can be viewed as an emergent property of a complex adaptive system. It is closely related to the concept of safety, which in such systems may arise from relationships, practices, and local adaptations rather than fixed procedures. This view is reflected in the distinction between Safety-I and Safety-II^28^. Safety-I, which has been the dominant model in healthcare, rests on a linear understanding of error, aiming to identify root causes of adverse events and prevent their recurrence. Safety-II shifts the focus toward how care succeeds, emphasising the value of understanding everyday work and learning from all forms of variation, not only from deviations. Most trauma quality improvement programs involving case review are structured around a Safety-I logic: defining clinical standards, detecting deviations, and seeking corrective action. While our intervention was similarly framed, using audit filter violations to select cases, the mechanisms through which improvement occurred were not tied to isolated errors. Instead, they emerged through mechanisms of system-level situational awareness, shared understanding, and the capacity to navigate the system.

### Limitations

This study was conducted in two urban hospitals in India, and while the mechanisms we identified may reflect common responses to uncertainty and fragmentation in health systems, their transferability is shaped by local context, available resources, and organisational culture. We use the term “mechanism” in an interpretive sense to describe emergent patterns of interaction and adaptation that emerge when individuals engage with the conditions and constraints of the system, rather than fixed causal pathways. These mechanisms are not exhaustive; they are plausible patterns that emerged under the conditions of this intervention, but other mechanisms may also contribute to improvement. Most participants were senior clinicians involved in the review process, which may have influenced the findings toward those with positional authority and the capacity to act on insights. As such, the mechanisms described may not reflect the experiences or constraints of more junior staff.

### Implications and future work

By identifying mechanisms that contribute to quality improvement in complex adaptive systems, we have gained a better understanding of how these programs may mediate their effect. Understanding how to improve quality improvement is key to addressing the known quality gap in global care, particularly for trauma. While these mechanisms emerged during structured case review, we believe they may represent a broader human response to working in complex systems with high uncertainty. The findings highlight that health systems are made and sustained by the people within them, whose relationships, judgement, and capacity to adapt are central to improvement. Supporting this human work, alongside structural change, is essential. Although these results are from university hospitals in urban India, the underlying principles may be applicable in other contexts. Future work should examine the presence of the identified mechanisms, as well as others not observed here, the conditions that enable them, and how programmes can sustain engagement over time.

## Conclusion

In this study, we identified four mechanisms that contributed to improvement during multidisciplinary case review: system-level awareness, shared understanding, navigating systems of power, and ethical sensitisation. These mechanisms emerged when a simple structure for case review was introduced. While our findings are grounded in a specific setting, we propose that the mechanisms identified may reflect a broader human response to working with uncertainty and interdependence in a complex adaptive system. Further research is needed to explore whether these mechanisms arise in other contexts, what conditions support their emergence, and how best to support healthcare providers within complex systems.

## Data Availability

Data (The transcripts form interviews) will not be available outside the research team due to the risk of identifying participants.

